# Comparative analysis of WGS and WES for genetic diagnosis in a pediatric Albanian population

**DOI:** 10.1101/2025.07.24.25332056

**Authors:** P. Cullufi, M. Tabaku, A. Skrahin, S. Tomori, V. Velmishi, E. Dervishi, G. Hoxha, A. Gjikopulli, E. Troja, A. Tako, V. Skrahina, A. Rolfs

## Abstract

**Background:** Rare diseases affect over 300 million people worldwide, with more than half of these cases presenting in childhood. Despite advances in genomic technologies, significant diagnostic delays remain, particularly in underrepresented populations. Although whole-genome sequencing is expected to be more effective than whole-exome sequencing, there are currently no direct patient-level comparisons from Albania.

**Methods:** We conducted a prospective, head-to-head comparison of whole-genome sequencing and whole-exome sequencing in 72 pediatric patients from Albania with suspected rare genetic disorders. Both technologies were applied in parallel using ISO-accredited pipelines. Variants were classified according to the ACMG guidelines and assessed for clinical relevance. Primary and secondary findings, as well as dual diagnoses, were recorded and compared.

**Results:** A molecular diagnosis was achieved in 72.2% of patients. Whole genome sequencing yielded diagnostic or secondary findings in 68.1% of cases, while whole exome sequencing identified primary diagnoses in 30.6% of cases. WGS made a primary diagnosis in 37.5% of cases, resolving complex or blended phenotypes and detecting variant types that were missed by WES, including deep intronic, regulatory and structural variants. Furthermore, WGS identified medically actionable secondary findings in 15.3% of patients, providing direct information for clinical management. Overall, WGS outperformed WES across variant classes and inheritance modes.

**Conclusion:** WGS demonstrated clear diagnostic superiority over WES in this pediatric cohort, particularly in identifying complex phenotypes and structural variants. These results suggest that WGS should be considered a primary diagnostic tool and highlight the importance of ensuring equitable access to genomic technologies for underrepresented populations.

## INTRODUCTION

Rare diseases (RDs) affect around 300 million people worldwide, with more than half of cases presenting in childhood, often within the first two years of life (1–2). These conditions are usually severe and affect multiple systems in the body, and they are also genetically heterogeneous, which poses significant diagnostic challenges. Clinical manifestations may be atypical, age-dependent or overlap between syndromes, which frequently results in delayed or missed diagnoses and a prolonged diagnostic odyssey (3– 6). On average, patients experience a delay of 5–7 years in receiving a diagnosis, often undergoing multiple inconclusive investigations in the process (7–8).

Traditional genetic testing approaches, such as chromosomal microarrays or targeted panels, have limited diagnostic value in genetically heterogeneous patient groups. The introduction of next-generation sequencing (NGS) has transformed diagnostics by enabling broad genomic interrogation. Whole-exome sequencing (WES), which targets the 1–2% of the genome that encodes proteins, is widely used due to its cost-effectiveness and clinical utility. Diagnostic yields in pediatric populations are reported to be between 25 and 50% (3–4, 9). Nevertheless, 40–70% of patients remain undiagnosed following WES, particularly those with atypical phenotypes or from populations with limited genomic data (10–11).

By contrast, whole genome sequencing (WGS) interrogates the entire genomic landscape, including both coding and non-coding regions. This enables the detection of a broader range of pathogenic variants, such as structural rearrangements, deep intronic changes and regulatory mutations (5, 12–13). Recent large-scale studies have demonstrated that WGS improves diagnostic rates by up to 60%, while also enhancing the detection of blended phenotypes, secondary findings, and actionable variants relevant to precision medicine (14, 15). The early adoption of WGS has been associated with reduced healthcare costs and a shorter time to diagnosis (16–19).

Despite these advantages, WGS implementation remains limited in low- and middle-income countries (LMICs), largely due to infrastructural, financial and workforce constraints (9, 20–21). The Albanian pediatric population is particularly underrepresented in genomic research, which limits variant interpretation and highlights the need for locally derived data. To address this, we present the first head-to-head comparison at the patient level of WGS and WES in an Albanian pediatric cohort. Our findings support recent evidence that WGS is superior for diagnosis in diverse pediatric populations (14,15). We assess the diagnostic yield, variant spectrum and clinical impact of both technologies, thereby contributing to the global effort to reduce diagnostic inequities and ensure equitable access to genomic medicine.

## MATERIAL AND METHODS

72 patients were enrolled in the study between 2020 and 2021. Conducted at the Department of Pediatrics at Mother Teresa University Centre, the study enrolled patients from all regions of Albania presenting with neurodevelopmental disorders, congenital anomalies, endocrine diseases, growth abnormalities, distinct physical phenotypes, atypical behaviours, or suspected genetic disorders of unknown etiology. Patients under the age of 18 were included in the study.

Informed consent for participation in the study, including the collection of blood and saliva samples (buccal swabs), the processing of personal data and the storage of biological material, was obtained from all patients or their legal guardians before enrolment. The study adhered to the ethical standards set by the National Ethics Committee and was conducted with approval number 303/15.

Saliva and blood samples from all 72 patients were sent to Laboratory 1, while dried blood spots from the same patients were sent to Laboratory 2. Laboratory 1 performed whole genome sequencing (WGS), while laboratory 2 performed whole exome sequencing (WES). While WGS and WES were performed in different laboratories, both adhered to ISO-accredited protocols with stringent quality control and depth metrics, ensuring data comparability.

### Whole Exome Sequencing (WES)

Venous blood was collected from each of the enrolled patients and their parents in the EDTA vacutainers. Genomic DNA was extracted using the Gentra Puregene Blood Extraction Kit (Qiagen, Germantown, Maryland, USA).

The experimental workflow of all exomes was performed at Lab 1 (Germany). Briefly, double-stranded DNA capture baits against approximately 36.5 Mb of the human coding exome, were used to enrich target exonic regions from fragmented genomic DNA with the Human Core Exome Plus kit (Twist, Bioscience), according to manufacturer’s instruction. The generated library was sequenced using an Illumina HiSeq 2500 (Vienna, Austria) sequencer with 101-bp paired end reads to obtain at least 20x coverage depth for >98% of the targeted bases. Ingenuity Variant Analysis software (2012 beta release; QIAGEN) was used to analyze the WES data. The investigation for relevant variants was focused on coding exons and flanking +/-20 intronic nucleotides of genes with clear genetype-phenotype evidence (based on OMIM® information). The standard nomenclature recommended by the Human Genome Variation Society was employed to number mutations.

### Whole Genome Sequencing (WGS)

The experimental workflow of all genomes was performed at Lab 2 (Germany). DNA samples were prepared using the TruSeq DNA Nano Library Prep Kit from Illumina. The libraries were pooled and sequenced with the 150-bp paired-end protocol on an Illumina platform to yield an average coverage depth of 30× for the nuclear genome. Raw read alignment to reference genome GRCH38 and variant calling, including single nucleotide substitutions (SNVs), small insertions/deletions (Indels), and structural variants (SVs) with default parameters, were performed using DRAGEN (version 3.10.4, Illumina). SNV and indel annotation were performed by Varvis (Limbus Medical Technologies GmbH; https://www.limbus-medtec.com/). Structural variants were annotated with ANNOTSV3.1 and the in-house structural variant database to obtain occurrence frequencies. Genetic variants are described after the Human Genome Variation Society recommendations (https://varnomen.hgvs.org/).

### Variant evaluation and interpretation

Only good-quality variants with a minimum of nine reads and an alternate allele frequency of at least 0.3% were considered. Candidate variants were evaluated with respect to their pathogenicity and causality and categorized following ACMG guidelines (22) using the five-tier classes: pathogenic (P), likely pathogenic (LP), variants of uncertain significance (VUS), likely benign (LB), and benign (B). Variants were assessed in a routine diagnostic setting. An initial analysis was restricted to genes that have a clear association with the participant’s phenotype using Human Phenotype Ontology nomenclature (https://hpo.jax.org/app/). The variants found in these genes were “the primary findings.” During this step, common standards were followed. Briefly, the following aspects were considered: the minor frequency of the allele in control databases (gnomAD) and internal database; the in silico pathogenicity prediction and potential impact on respective proteins; the known mechanism of the variant type-disease (missense, truncating, etc.); segregation in the family; and available external evidence: OMIM (https://www.omim.org/), ClinVar information, Mastermind, and genotype-phenotype correlation. Secondary findings (SF) that do not correlate with the provided phenotype(s) were recorded and reported according to ACMG recommendations version 3.1 (2022) for reporting of SFs in clinical exome and genome sequencing 1716, which was the current version during the study period.

### Sanger sequencing

All the filtered variants were validated by bi-directional Sanger sequencing. Sequencing was performed using Veriti 96-Well Fast Thermal Cycler and ABI Prism 3730 Genetic Analyzer (California, USA) Finch TV (1.4.0) and Seqtrace v0.9.0 software.

### Statistical analysis

Fisher’s exact test was used to compare the diagnostic yields of WGS and WES. A p-value of less than 0.05 was considered statistically significant. Given the exploratory nature of the study, no adjustments for multiple comparisons were applied.

## RESULTS

### Diagnostic yield and clinical contribution

In this clinically heterogeneous cohort of 72 pediatric patients with suspected genetic disorders, a molecular diagnosis was achieved in 52 individuals (72.2%) through WGS, WES or both. Specifically, WGS contributed to a primary diagnosis and/or secondary findings in 49 patients (68.1%), while WES identified the primary diagnosis in 22 patients (30.6%). All proportions are calculated based on the full cohort (n = 72), unless otherwise indicated. The cohort included patients with various clinical presentations, such as neurodevelopmental delay, congenital anomalies, and suspected metabolic syndromes.

### Comparing the diagnostic yield of WGS and WES

A diagnosis was obtained exclusively through WGS for 27 patients (37.5%). Nineteen patients (26.4%) had diagnostic variants detected by both technologies. WES identified the primary diagnosis in three patients (4.2%) exclusively; in these cases, WGS revealed only secondary findings. A further three patients (4.2%) remained without a primary diagnosis, but WGS identified actionable secondary findings in these cases (see Tables 1-5).

**Table 1.** Primary diagnosis detected only by WGS.

| Nr. | Patient ID | Gene | Variant (cDNA) | Protein Change | ACMG Class | Zygosity | MOI | Diagnosis |
| --- | --- | --- | --- | --- | --- | --- | --- | --- |
| 1 | P08 | G6PD | c.1360C>T | p.(Arg454Cys) | PV | Heterozygous | XL | G6PD |
|  |  | CUL7 | c.5047A>C | p.(Thr1683Pro) | VUS | Homozygous | AR | 3-M syndrome type 1 |
| 2 | P10 | CPLANE1 | c.7817T>A,<br>c.1819del | p.(Leu2606*)<br>p.(Tyr607Thrfs*6) | LP<br>LP | Compound heterozygote | AR | CLANE1<br>OFD6, JBTS17 |
| 3 | P11 | FGFR1 | c.83C>T | p.(Pro28Leu) | VUS | Heterozygous | AD | KS |
| 4 | P12 | CDKL5 | c.1616G>C | p.(Arg539Thr) | VUS | Heterozygous | XLD | DEE2 |
| 5 | P13 | EIF2AK2 | c.101G>A | p.(Gly34Glu) | VUS | Heterozygous | AD | LEUDEN |
| 6 | P14 | GNAS | c.3013C>A | p.(His1005Asn) | VUS | Heterozygous | AD | Disorders of GNAS Inactivation |
| 7 | 15 | SPTA1 | c.4844G>A, | p.(Ser1615Asn) | VUS | Heterozygous | AD | EL2 |
| 8 | P16 | CLTC | c.3218G>A, | p.(Arg1073Gln) | VUS | Heterozygous | AD | MRD56 |
| 9 | P17 | COL5A1 | c.4706C>T | p.(Pro1569Leu) | VUS | Heterozygous | AD | EDSCL1 |
| 10 | P18 | PRKCG | c.1381G>A | p.(Ala461Thr) | VUS | Heterozygous | AD | SCA14 |
| 11 | P19 | ABCC6 | c.3413G>A | p.(Arg1138Gln) | LP | Heterozygous | AD | PXE. forme fruste |
| 12 | P20 | WNT5A | c.422T>C | p.(Val141Ala) | LP | Heterozygous | AD | DRS1 |
| 13 | P21 | NIPBL | c.4344_4345d | p.(His1448Glnfs*30) | LP | Heterozygous | AD | CDLS1 |
| 14 | P22 | SMARCC2 | c.1863+3A>G | p.? | VUS | Heterozygous | AD | CSS8 |
| 15 | P23 | KCNT1 | c.3178-1G>C | p.? | LP | Heterozygous | AD | DEE14 |
| 16 | P24 | SPTBN1, | p.(Glu1986*) | NM_003128.3 | LP | Heterozygous | AD | DDISBA |
| 17 | P25 | RYR1 | c.4333_4336d | p.(Ser1445Alafs*16) | LP | Heterozygous | AD | RYR1- related disorders |
| 18 | P26 | PIK3R1 | c.1048A>G | p.(Thr350Ala) | VUS | Heterozygous | AD | SHORT syndrome |
| 19 | P27 | CACNA1A | c.6656A>C, | p.(His2219Pro) | VUS | Heterozygous | AD | DEE42 |
| 20 | P57 | NPRL3 | c.1270C>T | p.(Arg424*) | P | Heterozygous | AD | FFE type 3 |
| 21 | P62 | MYH3 | c.3137_3138delins<br>AC | p.(Arg1046His) | VUS | Heterozygous | AD | MYH3 related disorders |
| Nr. | Patient ID | Locus | Genomic position | Copy nr./ Type | ACMG class | Zigosity | MOI | Diagnosis |
| 22 | P35 | 16p11.2 | Chr16:<br>29,569,549-30,189,076 | 1/ Loss | LP | Heterozygous | AD | 16p11.2 recurrent microdeletion |
| 23 | P36 | 12p11.1p13.33 | Chr12:10501-34718907 | 3/Gain | LP | Heterozygous | AD | Trisomy 12p |
| 24 | P37 | Non determined |  |  |  |  | AD | large heterozygous deletion, disrupting the LCN1 gene |
| 25 | P38 | Non determined |  |  |  |  | AD | large heterozygous deletion comprising the entire ALAD gene |
| 26 | P39 | Xq26.3q28 | ChrX:<br>135,760,044<br>153,427,263 | 2/Gain | LP | Heterozygous | XL | Chromosome Xq26.3q28 duplication syndrome |
| 27 | P48 | Xp22.31 | X:6986366-7371841 | Gain | VUS | Heterozygous | XL | Ichthyosis |

Abbreviations: ACMG = American College of Medical Genetics and Genomics; PV = Pathogenic Variant; LP = Likely Pathogenic; VUS = Variant of Uncertain Significance; P = Pathogenic; MOI = Mode of Inheritance; AD = Autosomal Dominant; AR = Autosomal Recessive; XL = X-linked; XLD = X-linked Dominant

### Primary diagnoses identified exclusively by WGS

WGS uniquely resolved 27 cases that were undetected by WES (see Table 1). All structural or copy number variation (CNV) findings were technically validated via multiplex ligation-dependent probe amplification (MLPA) or array comparative genomic hybridisation (array CGH) when applicable. These representative cases highlight variant types that are only captured by WGS and are likely to be missed by WES due to limitations in probe design or analytical constraints. Examples include:

- Deep intronic splice-site variants in *SMARCC2* (P22) and *KCNT1* (P23).
- Poorly captured exonic variants, such as multiple nucleotide substitutions in *MYH3* (P62);
- Structural and copy number variants: - 16p11.2 microdeletion (P35); - ∼34.7 Mb duplication of chromosome 12p (P36); - Xq26.3–q28 duplication (P39); - Xp22.31 gain (P48).
- Compound heterozygous and regulatory variants in *CPLANE1* (P10);
- Novel or rare variants, including a frameshift in *NIPBL* (P21), a nonsense variant in *SPTBN1* (P24) and missense variants in *WNT5A* (P20) and *ABCC6* (P19).

Previously unreported deletions in *LCN1* (P37) and *ALAD* (P38) suggest novel gene– phenotype correlations that merit further investigation.

### Concordant findings between WGS and WES

Nineteen patients (26.4%) had diagnostic variants identified by both WGS and WES. Variant classification was fully concordant in 17 of these cases (see Table 2). Representative examples include:

**Table 2.**
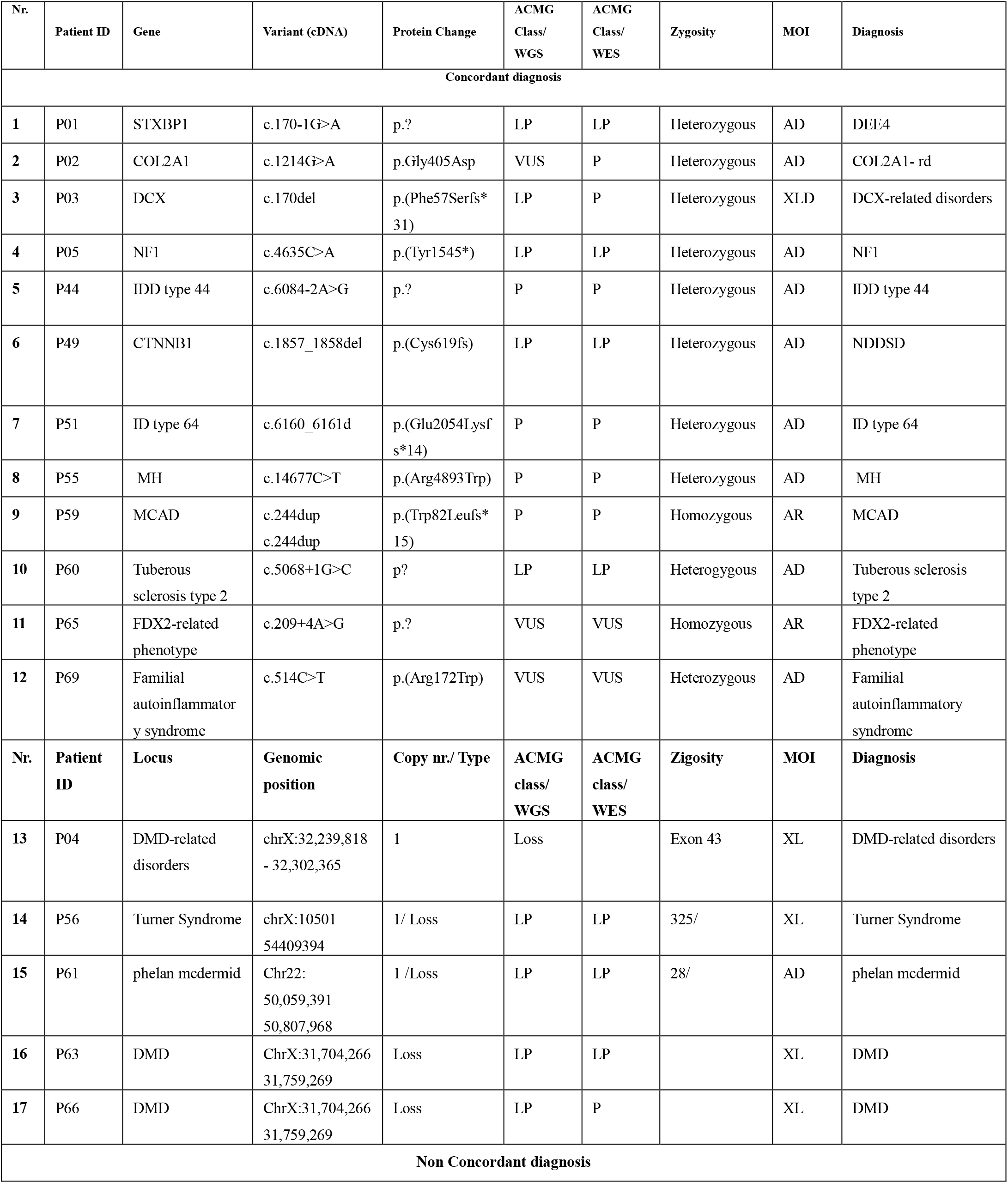
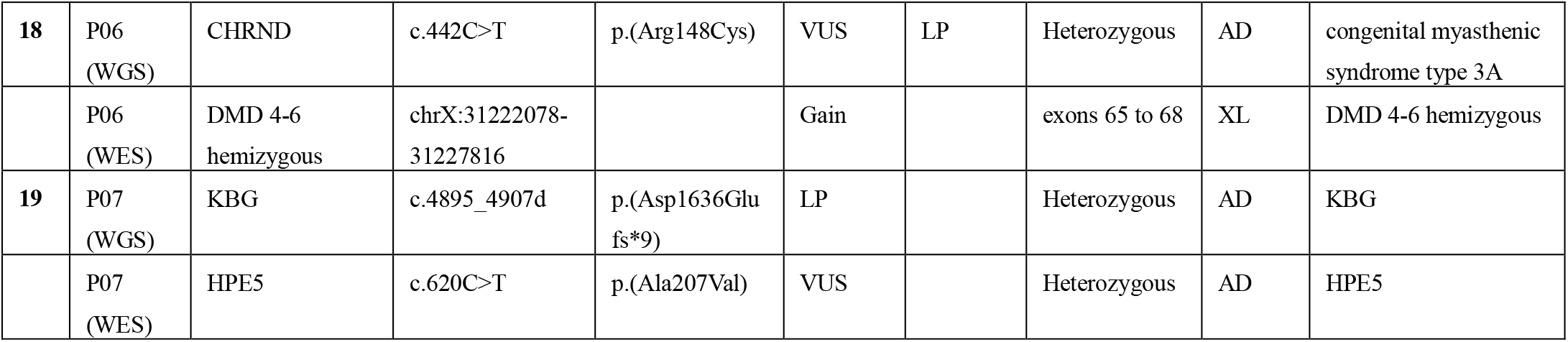
Primary dg detected by both WGS and WES.

- Canonical splice-site variants: *STXBP1* (P01) and *TSC2* (P60).
- Truncating variants: *DCX* (P03), *ZNF292* (P51), *CTNNB1* (P49), and *NF1* (P05).
- Missense variants: *TRIO* (P44) and *RYR1* (P55).
- A homozygous loss-of-function variant in *ACADM* (P59).
- Intragenic deletions in *DMD* (P04, P63, and P66).
- Large structural variants (SVs): a 54 Mb X-chromosome deletion (Turner syndrome, P56) and a 749 Kb deletion at 22q13.33 (Phelan-McDermid syndrome, P61).

Minor discrepancies in variant classification (e.g. likely pathogenic vs. VUS) were observed.

### Discordant findings

P06: WES with copy number variation (CNV) analysis identified a *DMD* duplication (exons 65–67), which was confirmed by multiplex ligation-dependent probe amplification (MLPA). However, WGS failed to detect this and instead reported an unrelated *CHRND* variant. P07: WGS detected a likely pathogenic variant in *ANKRD11*, which is consistent with KBG syndrome. WES identified a de novo VUS in *ZIC2* that lacked phenotypic relevance.

### Primary diagnostic findings from WES only

WES resolved three cases exclusively (Table 3):

**Table 3.** WES based primary diagnosis.

| Nr. | Patient ID | Gene | Variant (cDNA) | Protein Change | ACMG Class | Zygosity | MOI | Diagnosis |
| --- | --- | --- | --- | --- | --- | --- | --- | --- |
| **1** | P32 | ASXL1<br>CBL | c.1900_1922<br>del<br>c.1259G>A | p.(Glu635Argfs*1<br>5)<br>p.(Arg420Gln) | LP<br>LP | Heterozygous<br>Heterozygous | AD | MDS<br>MPD |
| **2** | P33 | FDX2, | c.209+4A>G | p? | VUS | Homozygous | AR | MEOAL |
| Nr. | Patient ID | Locus | Genomic position | Copy nr./ Type | ACMG class | Zigosity | MOI | Diagnosis |
| **3** | P30 | 22q13.32q<br>13.33 | 48660023_5<br>1197766 | 1 | Loss | Heterozygous | AD | Chromosome 22q13.3 deletion syndrome (Phelan-Mcdermid syndrome) |

- P30: terminal deletion at 22q13.32–q13.33 (Phelan–McDermid syndrome);
- P32: two heterozygous likely pathogenic variants in *ASXL1* and *CBL* (MDS/MPD overlap);
- P33: Homozygous intronic variant of uncertain significance (VUS) in *FDX2*, compatible with an MEOAL-like phenotype.

WGS only contributed secondary findings in these patients.

### Secondary findings identified exclusively by WGS

WGS revealed medically actionable secondary findings in 11 patients (15.3%), all of which were undetected by WES (see Table 4). These include:

**Tab.4.**
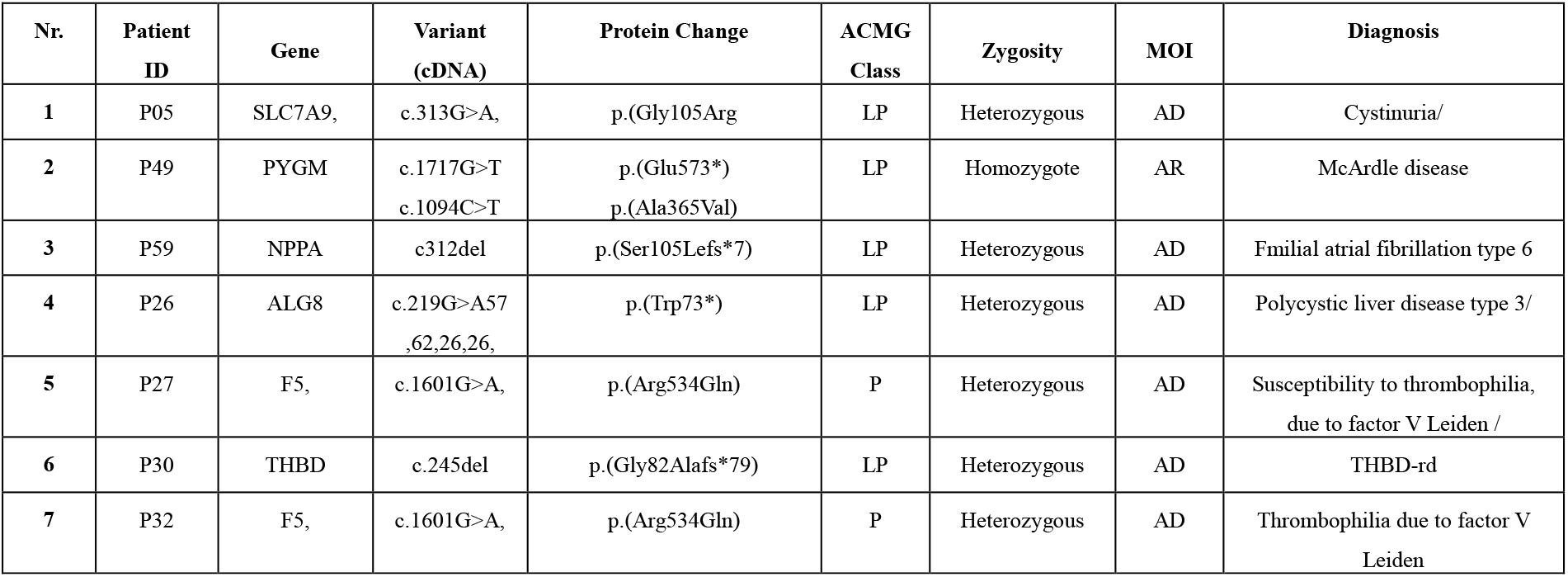
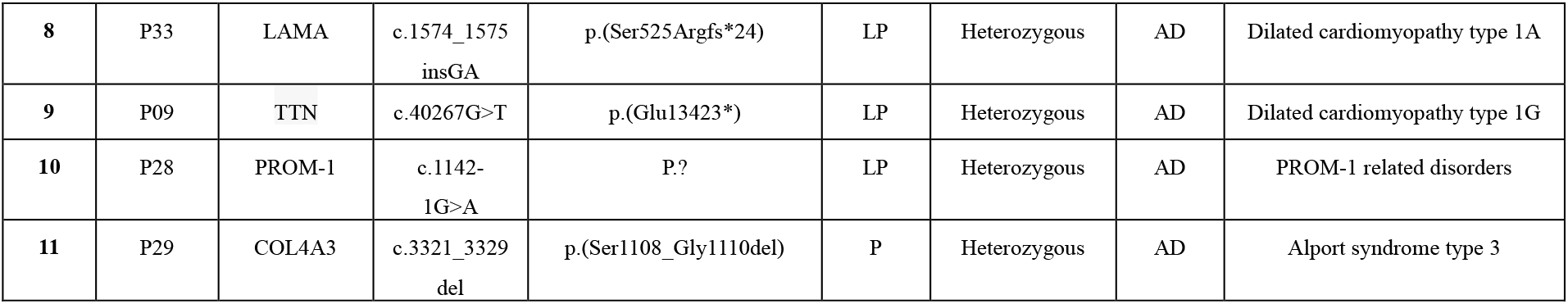
Secondary findings: patients with WGS-based primary diagnoses (1-5), patients with WES-based primary diagnoses (6-8), patients with no primary diagnosis (9-11).

- Patients with no primary diagnosis: P09 (*TTN*), P28 (*PROM1*), and P29 (*COL4A3*).
- Patients with a primary diagnosis identified by WGS: P05 (*SLC7A9*), P26 (*ALG8*), P27 (*F5*), P49 (*PYGM*), and P59 (*NPPA*).
- Patients with a primary diagnosis identified by WES: P30 (*THBD*), P32 (*F5*), and P33 (*LAMA2*).

These findings prompted targeted surveillance, specialist referrals and cascade testing, thereby reinforcing the preventive and clinical utility of WGS.

### Dual molecular diagnoses (Table 5)

WGS uniquely revealed dual molecular diagnoses in three patients: P05: neurofibromatosis type 1 with features of Ververi-Brady syndrome (*NF1* + *QRICH1*); P10: compound heterozygous *CPLANE1* variants combined with a 22q11.2 deletion; and P08: a *G6PD* missense variant together with a homozygous *CUL7* variant, suggestive of 3-M syndrome. WES revealed a dual diagnosis in one patient (P32) with variants in both *ASXL1* and *CBL*, which are consistent with myelodysplastic/myeloproliferative neoplasm (MDS/MPN) overlap syndrome.

**Tab.5.** Dual diagnosis by WGS (P05, P08, P10) and WES (P30)

| Nr. | Patient ID | Gene | Variant (cDNA) | Protein Change | ACMG Class | Zygosity | MOI | Diagnosis |
| --- | --- | --- | --- | --- | --- | --- | --- | --- |
| 1 | P05 | QRICH1 | c.577C>T | p.(Gln193*) | LP | Heterozygous | AD | VERBRAS |
|  |  | NF1 | c.4635C>A | p.(Tyr1545*) | LP | Heterozygous | AD | NF1 |
| 2 | P08 | G6PD | c.1360C>T | p.(Arg454Cys) | PV | Heterozygous | XL | G6PD |
|  |  | CUL7 | c.5047A>C | p.(Thr1683Pro) | VUS | Homozygous | AR | 3-M syndrome type 1 |
| 3 | P10 | CPLANE1 | c.7817T>A<br>,<br>c.1819del | p.(Leu2606*)<br>p.(Tyr607Thrfs*6) | LP<br>LP | Compound heterozygote | AR | CLANE1<br>OFD6, JBTS17 |
| Nr. | Patient ID | Locus | Genomic position | Copy nr./ Type | ACMG class | Zigosity | MOI | Diagnosis |
| 3 | P10 | 22q11.21 | Chr22:<br>18,811,852-19,026,179 | 1 | VUS | Heterozygous | AD | 22q11.2DS |
|  | Patient ID | Gene | Variant (cDNA) | Protein Change | ACMG Class | Zygosity | MOI | Diagnosis |
| 4 | P30 | ASXL1 | c.1900_192<br>2del | p.(Glu635Argfs*1<br>5) | LP | Heterozygous | AD | MDS |
|  |  | CBL | c.1259G>A | p.(Arg420Gln) | LP | Heterozygous | AD | MPD |

**Table 6.** Variant Spectrum.

| Variant Type | WGS Nr. | WES Nr. |
| --- | --- | --- |
| Missense | 22 | 6 |
| Frameshift | 14 | 5 |
| Nonsense | 8 | 1 |
| Splice-site | 7 | 4 |
| Structural Variants | 12 | 7 |
| Total nr | 63 | 23 |

### Variant spectrum and clinical classification

WGS identified 63 clinically significant variants (81% single nucleotide variants (SNVs), 19% structural variants (SVs)), whereas WES identified 23 (70% SNVs, 30% SVs). WGS demonstrated greater sensitivity across all variant types, particularly for missense, frameshift, nonsense, splice-site and structural variants (Table 7).

**Table 7.** ACMG classification of detected variants.

| Classification | WGS |  | WES |  |
| --- | --- | --- | --- | --- |
|  | Nr | % | Nr. | % |
| **Pathogenic (P)** | 10 | 15.9 | 9 | 39.1 |
| **Likely Pathogenic (LP)** | 31 | 49.1 | 9 | 39.1 |
| **VUS ( Variant of Uncertain Significance)** | 19 | 30.2 | 4 | 17.4 |
| **Undetermined** | 3 | 4.8 | 1 | 4.4 |
| **Total nr.** | 63 | 100 | 23 | 100 |

WGS also yielded more likely pathogenic variants and clinically relevant variants of uncertain significance than WES (Table 8). Although both platforms detected a similar number of pathogenic variants, WGS demonstrated broader coverage and variant diversity, particularly in non-coding, structural and multi-nucleotide regions.

**Table 8.**
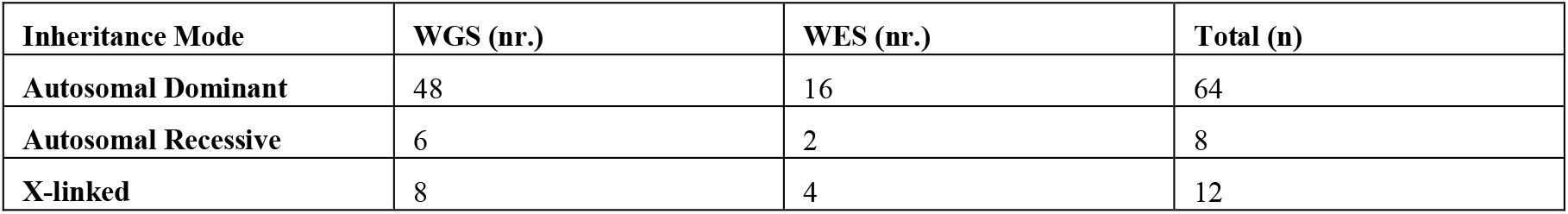
Inheritance patterns of diagnosed cases.

### Inheritance pattern analysis

Among the 52 patients with confirmed molecular diagnoses, including those with dual diagnoses and secondary findings, autosomal dominant inheritance was the most common (48 cases by WGS and 16 cases by WES), followed by X-linked inheritance (eight cases by WGS and four cases by WES) and autosomal recessive inheritance (six cases by WGS and two cases by WES) (see Table 9).

## DISCUSSION

Building on previous work from high-income countries, this study presents the first parallel, patient-level comparison of whole genome sequencing and whole exome sequencing in an Albanian pediatric cohort. This addresses a critical gap in the literature on underrepresented populations, as recently highlighted in meta-analyses (14, 15). By applying both technologies simultaneously in 72 children with suspected rare genetic disorders, we were able to directly compare their diagnostic yield, variant spectrum and clinical impact in a healthcare setting with limited resources.

### Diagnostic yield and variant detection

In our cohort, a molecular diagnosis was achieved in 72.2% of patients. WGS contributed to a diagnosis or secondary findings in 68.1% of cases, while WES identified a primary diagnosis in 30.6% of cases. Notably, WGS resolved 37.5% of cases that remained undiagnosed by WES alone. A further 26.4% of patients received diagnoses through both technologies. While previous large-scale studies have reported comparable yields for WGS and WES in mixed cohorts, our findings demonstrate a notably higher yield for WGS (68.1%). The high diagnostic yield in our study likely reflects methodological and technological strengths. Broad inclusion criteria and expert-driven standardised phenotyping enhanced genotype–phenotype correlation, while the superior variant detection capacity of WGS and targeted case selection based on phenotypic complexity significantly boosted diagnostic success.

The diagnostic advantage of WGS was particularly evident in patients with non-specific neurodevelopmental presentations, for which traditional testing is often insufficient. WGS also proved crucial in resolving complex phenotypes, as demonstrated by dual molecular diagnoses in patients P05 (*NF1* + *QRICH1*), P08 (*G6PD* + *CUL7*), and P10 (*CPLANE1* + 22q11.2 deletion) all of which remained unresolved with WES alone. These findings are consistent with previous estimates that up to 5% of individuals with a genomic diagnosis may harbor more than one Mendelian condition (15). This highlights the usefulness of WGS in identifying multilocus or blended phenotypes, which are a diagnostically challenging category that is becoming increasingly recognized in clinical genetics.

### Breadth and complexity of variants

WGS identified 63 clinically relevant variants across the cohort, including single-nucleotide variants (SNVs), small insertions/deletions (indels) and large structural variants (SVs). In contrast, WES detected only 23 variants of this type. WGS demonstrated superior sensitivity across all major variant classes (5–6, 23–24). Several WGS-unique variants were located in regions with limited WES coverage, including deep intronic changes and multi-nucleotide substitutions, which is consistent with earlier reports (12–13). These variants were found in *MYH3* (P62), *SMARCC2* (P22), *KCNT1* (P23), *NIPBL* (P21), *SPTBN1* (P24), *WNT5A* (P20), *ABCC6* (P19), and *RARS1* (P44). Structural variants that were only identified by WGS included a copy number variation (CNV) in *STXBP1* (P41), a ∼34 Mb duplication at Xq26.3–q28 (P39), a 16p11.2 deletion (P35), and partial trisomy of 12p (P36). Most of these were not confidently resolved by WES. Furthermore, WGS revealed previously unreported deletions in *LCN1* and *ALAD*, suggesting potential novel gene–phenotype associations. Although preliminary, these findings warrant further functional studies and familial segregation analysis to establish causality. They highlight the hypothesis-generating capacity of WGS and its role in broadening the recognised phenotypic spectrum of rare diseases (15, 25).

Previous studies have estimated that 20–25% of diagnoses made via WGS involve variant types that are typically missed by WES (5, 26–27), a trend that is corroborated by our data. However, as demonstrated by the missed *DMD* duplication in P06, even WGS can fail to detect certain structural variants (SVs), particularly in GC-rich or highly repetitive regions. This highlights the need for continuous refinement of SV detection algorithms.

WGS provided diagnoses across all inheritance modes, including complex and blended cases, demonstrating its applicability in genetically heterogeneous pediatric populations.

### Secondary findings and preventive health value

WGS uncovered medically actionable secondary findings in 15.3% of patients, none of which were identified by WES. This rate exceeds the 1–5% secondary finding rates typically reported in adult populations (28) and is possibly due to the younger age of this cohort and the broader variant analysis. These findings prompted follow-up interventions in multiple patients. Examples included: Patients P09 (*TTN*) and P33 (*LAMA2*) were referred for cardiac surveillance: P29 (*COL4A3*) referred for a nephrology consultation: P28 (*PROM1*) referred for a retinal evaluation: P27 and P32 (*F5*) referred for thromboprophylaxis counselling. Other relevant findings included an *SLC7A9* variant in P05 (cystinuria); a *PYGM* variant in P49 (McArdle disease susceptibility); and a *LAMA2* variant in a child with *ATP7A*-related Menkes disease. The latter finding led to early cardiac monitoring. All returns were handled according to ACMG v3.1 and ethical standards, including the Declaration of Helsinki (19, 28–29). These findings support the growing role of WGS in diagnosis, anticipatory care, and public health, particularly in pediatrics (30–31). Identifying secondary findings promotes proactive risk stratification and early intervention, representing a shift towards preventive genomic medicine.

### Broader implications

The superior diagnostic capacity of WGS translates into clinically meaningful benefits. It enables the resolution of complex cases and can directly guide medical management in a significant proportion of patients (8, 19, 23). For families experiencing prolonged diagnostic odysseys, the early implementation of WGS can provide definitive molecular diagnoses and facilitate timely access to personalized care and reproductive counselling. Although certain technical limitations remain, such as reduced sensitivity for specific structural variants in GC-rich regions, its overall performance aligns with current recommendations that advocate WGS as a first-tier diagnostic modality in genetically heterogeneous or unresolved cases (11, 15, 19, 32, 33).

However, accurate variant interpretation requires extensive manual curation, especially given the limited representation of Southeast European populations in widely used genomic reference databases such as gnomAD. The high proportion of rare and novel variants observed in our cohort highlights the urgent need for region-specific genomic data and collaborative, multi-institutional efforts to enhance variant annotation and classification frameworks.

### Study limitations

The modest sample size (n = 72) and single-center design may limit its generalizability. The composition of the cohort, which was enriched for neurodevelopmental and multisystem phenotypes, may have influenced the diagnostic yield. Furthermore, WGS and WES were performed in different laboratories, which could have introduced variability in sequencing platforms, coverage and bioinformatics pipelines.

No reanalysis of variants was performed, which may have affected the resolution of certain VUSs. Furthermore, interpretation of VUSs remains particularly challenging in underrepresented populations due to limited reference data. Although a formal cost-effectiveness analysis was not included, future work will incorporate clinical-economic modelling tailored to emerging healthcare systems (33). These considerations highlight the importance of regional variant databases and international collaboration in supporting the accurate interpretation of genomic data and ensuring equitable access to genomic medicine.

Despite its limitations, this study provides essential pilot data to inform national rare disease strategies and the inclusion of genomic sequencing in pediatric diagnostic algorithms.

## Conclusion

This comparative analysis shows that whole-genome sequencing provides greater diagnostic sensitivity and detects a wider range of variants than whole-exome sequencing when evaluating pediatric patients with suspected rare genetic disorders. Our findings, derived from an Albanian pediatric cohort, provide novel real-world evidence in support of the clinical utility of WGS in underrepresented populations.

WGS outperforms WES, particularly in the detection of structural variants, blended phenotypes, and clinically actionable findings, and therefore emerges as the preferred first-line approach for diagnostically challenging cases. Integrating WGS into clinical workflows has the potential to substantially improve diagnostic accuracy and accelerate access to precision care.

## Data Availability

All data produced in the present study are available upon reasonable request to the authors

